# Feasibility of Quantifying Mild Cognitive Impairment Using Multi-Modal Wearable Sensors in a Kitchen Environment

**DOI:** 10.1101/2025.05.24.25328107

**Authors:** Bonwoo Koo, Ibrahim Bilau, Amy D. Rodriguez, Eunhwa Yang, Hyeokhyen Kwon

**Author notes:** Joint Senior Authors (Eunhwa Yang, Hyeokhyen Kwon). Author to whom any correspondence should be addressed,.

## Abstract

Behavioral sensing using wearables has emerged as a valuable tool for screening of neurodegenerative conditions, including Mild Cognitive Impairment (MCI). Existing work has primarily focused on using wearables to quantify walking patterns in individuals with MCI, typically in controlled environments. On the other hand, the human activity recognition community has been actively studying methods to quantify kitchen activities, which are instrumental activities of daily living (IADLs). Previous studies have reported deficits in visuospatial navigation among individuals with MCI, which may affect functional independence in the kitchen. This work studies the use of wrist and eye-tracking wearable sensors to quantify kitchen activities in individuals with MCI. We collected multimodal datasets from 18 older adults (11 with MCI and 7 with normal cognition) while preparing a yogurt bowl. Our multimodal analysis model could classify the older adults with MCI and those with normal cognition with a 76% F1 score. The feature importance analysis showed an association between weaker upper-limb motor function and delayed eye movements and cognitive decline, consistent with previous findings in MCI research from lab studies. This pilot study demonstrates the feasibility of monitoring behavior markers of MCI in daily living settings and calls for further studies with larger-scale validation in individuals’ home environments.

## 1 Introduction

Mild Cognitive Impairment (MCI) is a precursor to Alzheimer’s disease and related dementia (ADRD). Recent epidemiological data indicate a significant upward trend in its global impact, with prevalence rising from 15.56% in 2022 [1] to 19.7% in 2023 [2]. Given that individuals with MCI increasingly struggle with complex everyday tasks, timely intervention is paramount for preserving functional independence [3, 4]. While the Montreal Cognitive Assessment (MoCA) [5] and Clinical Dementia Rating (CDR) [6] are frequently used to screen for MCI, these tools often fail to detect subtle and longitudinal progression [7, 8]. More importantly, they do not capture how MCI affects individuals’ cognitive function in daily living settings. Individuals with MCI often exhibit reduced working memory capacity, slower processing speed, and difficulties with visuospatial navigation, which significantly increase cognitive load during the sequential operations required in instrumental activities of daily living (IADLs), like kitchen activities [9]. This limitation underscores the need for new techniques to continuously monitor changes in the functional independence of individuals living with MCI in everyday settings.

Wearable sensors serve as a promising tool to measure physical activity and physiological signals continuously, providing a new opportunity to monitor an individual’s functions in daily living [10, 11, 12]. Previously, researchers have utilized diverse sensor placements, such as wrist-, waist-, or hip-worn accelerometers, to capture gait and turning activities indicative of cognitive decline [13, 14, 15]. For instance, Shi *et al*. [16] demonstrated that movement patterns captured via triaxial accelerometers on both the wrist and hip in real-world settings serve as strong predictors of cognitive impairment. Beyond physical motion, the integration of physiological and environmental signals has provided digital markers associated with the early stages of neurodegeneration. Saif *et al*. [17] identified disrupted sleep and abnormal heart rate variability (HRV) as early digital biomarkers of AD, while Sakal et al. [18] combined daily activity, sleep, and light exposure metrics to predict cognitive performance.

Recent advancements have further leveraged machine learning frameworks to quantify behavior features associated with dementia using wearable sensors. The CogAx framework, for example, used contrastive learning to simultaneously predict CDR scores and daily activities using Inertial Measurement Unit (IMU) sensors [19]. Similarly, wearable IMU sensing has been explored as a novel marker for monitoring real-time cognitive focus during daily interactions [20]. Moving toward the recognition of functional independence in home settings, Cao *et al*. [21] successfully monitored cognitive health by classifying specific handwashing activities using wrist-worn IMUs. Other than IMU sensors, smart glasses equipped with eye-tracking sensors were successfully used to capture oculomotor patterns, such as pupil dilation, gaze patterns, and fixation stability, which correlate significantly with cognitive decline [22, 23]. Much of this research has focused on identifying visual attention patterns. Davis and Sikorskii [24] identified distinct wayfinding behaviors in virtual simulations, while Haque *et al*. [25] successfully detected patients with MCI by analyzing fixation-based metrics such as the frequency, duration, and spatial distribution of gaze on specific regions of interest. Beyond mapping these general gaze distributions, the high temporal resolution of eye-tracking signals further enabled an objective assessment of neurocognitive function through the precise evaluation of saccadic eye movements [26, 27]. These saccadic metrics provided deeper insights into inhibitory control and processing speed, as demonstrated by Chehrehnegar *et al*. [28] and Zhang *et al*. [29], which found that individuals with MCI exhibit characteristic abnormalities, such as increased error rates, prolonged latencies, and diminished error-correction capabilities. Despite these clinical advancements, the majority of existing research remains limited to stationary, desktop monitor-based eye-tracking systems [30, 31, 32, 33], leaving a significant gap in the investigation of portable eye-tracking glasses within complex, naturalistic settings. While wearable technologies have shown promise in assessing cognitive decline, many studies are conducted in controlled environments [34, 14]. Thus, there remains a need to validate these systems within the complex, multi-step behavioral sequences characteristic of naturalistic home activities, such as meal preparation.

Previously, human activity recognition (HAR) research has been actively conducted in the kitchen, using wearable or ambient sensors [35, 36, 37, 38] to successfully recognize complex activities in kitchen environments [39, 40, 41]. Preparing a meal is a natural proxy for assessing functional independence [42], as older adults spend 80% to 90% of their time at home, where meal preparation activity can potentially provide digital markers associated with broader cognitive decline [43, 44, 45]. Cooking tasks require the orchestration of complex, sequential operations that simultaneously engage executive functions, attention, and memory within a goal-oriented timeframe [46, 47]. This multi-dimensional cognitive demand makes meal preparation particularly sensitive to the visuospatial navigation deficits frequently observed in individuals with MCI [48, 49]. Consequently, the inherent complexity of kitchen activities provides a unique opportunity to employ sensor-based technologies to capture subtle yet important behavioral markers of cognitive decline within naturalistic settings. Beyond its screening utility, the therapeutic potential of cooking in stimulating executive functions [46] further underscores the importance of monitoring these activities in daily life.

For HAR in a kitchen environment, vision-based sensors, particularly RGB cameras, have long been the primary modality. Early studies utilized multi-camera setups in laboratory environments to capture discrete actions such as pouring and mixing [37, 50], while more recent work has leveraged egocentric video to record unscripted tasks in home settings [40]. Despite their ability to capture rich contextual details, vision-based approaches often encounter significant barriers regarding scalability, environmental robustness, and, most critically, user privacy [51]. To address these limitations, wearable sensors, such as accelerometers and gyroscopes, have emerged as a compelling alternative. Wearable sensors directly capture kinematic data from the user’s movements while inherently maintaining privacy. Furthermore, their ability to provide continuous, long-term monitoring in diverse environments makes them particularly suitable for identifying the longitudinal behavioral shifts associated with cognitive impairment. Early efforts to capture cooking activities focused on establishing comprehensive multimodal frameworks, such as CMU-MMAC [52], which integrated body-worn IMUs with external audio-visual sensors. Subsequent research, including the OPPORTUNITY dataset [36], used body-mounted IMU configurations to monitor daily routines like breakfast preparation and cleanup. The KDDI smartwatch dataset [35] significantly extended the granularity of activity recognition by capturing 74 distinct kitchen actions using commercial smartwatches in naturalistic settings. Most recently, ActionSense [39] has introduced an extensive multimodal wearable suite, combining eye-tracking, muscle activity, and tactile sensing, for the analysis of complex kitchen tasks. Parallel to these advances in wearable research, recent vision-based frameworks such as CHEF-VL have demonstrated the reasoning capabilities of Vision-Language Models (VLMs) to detect not only discrete actions but also higher-level cognitive sequencing errors within standardized cooking tasks [53]. Building upon the previous research, the present study employs eye-tracking devices and wrist-worn sensors to quantify physiological eye movement and physical motion to detect cognitive impairment in a naturalistic kitchen environment, which remains unexplored.

In this study, we assessed the feasibility of detecting cognitive decline using multimodal wearable sensors while older adults with normal cognition (NC) and those with MCI performed meal preparation tasks. Our study was conducted in a semi-naturalistic kitchen environment originally designed to support cooking-based therapeutic activities, moving beyond simplistic lab-based paradigms and providing a natural environment for assessing physiological and behavioral correlates of normal cognition and MCI. Participants’ behaviors were recorded using wrist sensors and eye-tracking glasses. Our contributions include:

- *i)* The first multimodal dataset of kitchen activities was captured from 18 age-matched older adults (N=11 MCI, N=7 NC) in a semi-naturalistic kitchen environment.
- *ii)* A HAR-based screening system that identifies MCI through complex behavioral patterns during kitchen activities.
- *iii)* Identification of physical and physiological features obtained from wearable devices, such as movement intensity and saccadic dynamics, as objective markers for cognitive decline during kitchen activities.

## 2 Materials and Methods

### 2.1 Ethics Statement

This study recruited participants with MCI and NC with the approval from the Institutional Review Board (IRB) at Georgia Institute of Technology (IRB #2025-859) and Emory University (IRB #2023P006274), which was approved on November 15th, 2023. Participants provided written informed consent to participating in this study. The study period was between October, 2023 and November, 2024.

### 2.2 Participants

Table 1 provides an overview of participants’ characteristics. A total of 18 participants were recruited for this study through Charlie and Harriet Shaffer Cognitive Empowerment Program (CEP) [54] at Emory University. The cohort consisted of 11 participants clinically diagnosed with MCI (mean age: 73.5 ±4.0 years) at Emory’s Cognitive Neurology Clinic and 7 individuals with NC (mean age: 69.1 ±2.8 years). The diagnosis of MCI was established by experienced neurologists based on standardized clinical criteria, National Alzheimer’s Coordinating Center (NACC) Uniform Data Set (UDS) Version 3 [55], while also including (1) subjective cognitive complaints reported by the participant or a reliable informant, (2) objective evidence of cognitive decline relative to age-matched norms through cognitive testing, and (3) relatively preserved independence in IADLs. Participants with MCI had a mean MoCA score of 22.5 ± 3.0. NC participants were primarily recruited as care partners of our participants with MCI. Although MoCA scores were not collected for the NC group, these individuals reported full functional independence in complex daily environments. Furthermore, all NC participants confirmed the absence of cognitive impairment or neurological disorders by self-report. Specifically, this recruitment framework and the participant cohort from the CEP have been validated through other studies investigating context-aware assistive technologies and the impact of environmental design on cognitive load for older adults with MCI and NC, ensuring the clinical consistency and reliability of our control cohort [56, 9].

**Table 1:**
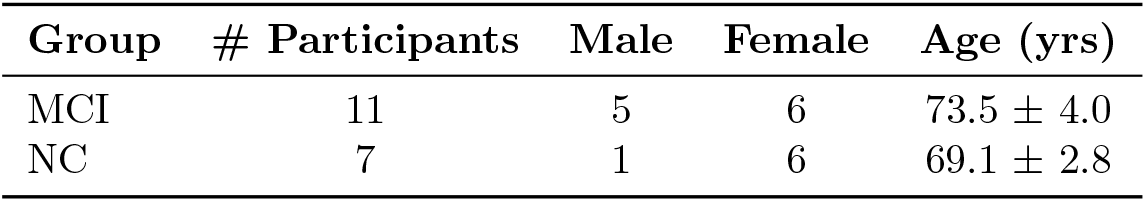
Participant Demographics.

| Group | # Participants | Male | Female | Age (yrs) |
| --- | --- | --- | --- | --- |
| MCI | 11 | 5 | 6 | $73.5 \pm 4.0$ |
| NC | 7 | 1 | 6 | $69.1 \pm 2.8$ |

### 2.3 Experimental Design: Yogurt Parfait Preparation Task

The experimental protocol involved a yogurt parfait preparation task, conducted in the kitchen space at the CEP (Figure 1). In this feasibility study, to ensure experimental consistency, all ingredients and utensils were organized in a standardized spatial configuration within the cabinets and refrigerator, as illustrated in the elevation and layout plans in Figure 2. The yogurt parfait preparation task was selected to balance participant safety with the need for high cognitive and motor demand sensitivity. By avoiding heat and sharp implements (e.g., knives), the task minimized physical risk to vulnerable participants [57]. The task consisted of 26 discrete steps (Table 2) designed to engage complex sequencing, visuospatial navigation, and motor coordination, reflecting standard IADLs per the Katz and Lawton scales [58]. While tasks are sufficiently brief to prevent fatigue, they allow for the detection of subtle deficits, such as delayed saccades or sequencing errors, consistent with therapeutic cooking paradigms [46]. Participants received both oral and written instructions at the start of the session. A printed 26-step recipe remained accessible on the countertop throughout the task to serve as a reference. All participants completed the task unassisted, and no steps were skipped. The protocol required participants to return used ingredients or utensils to their original locations (e.g., cabinet or refrigerator) before proceeding, ensuring the capture of naturalistic behavioral variations and the transition between two primary activity classes: *item retrieval* and *food preparation* (Table 2). Further technical details regarding the standardized 26-step yogurt parfait recipe, including the specific spatial arrangement of ingredients and utensils within the instrumented kitchen, are documented in Bilau et al. [9].

**Figure 1:**
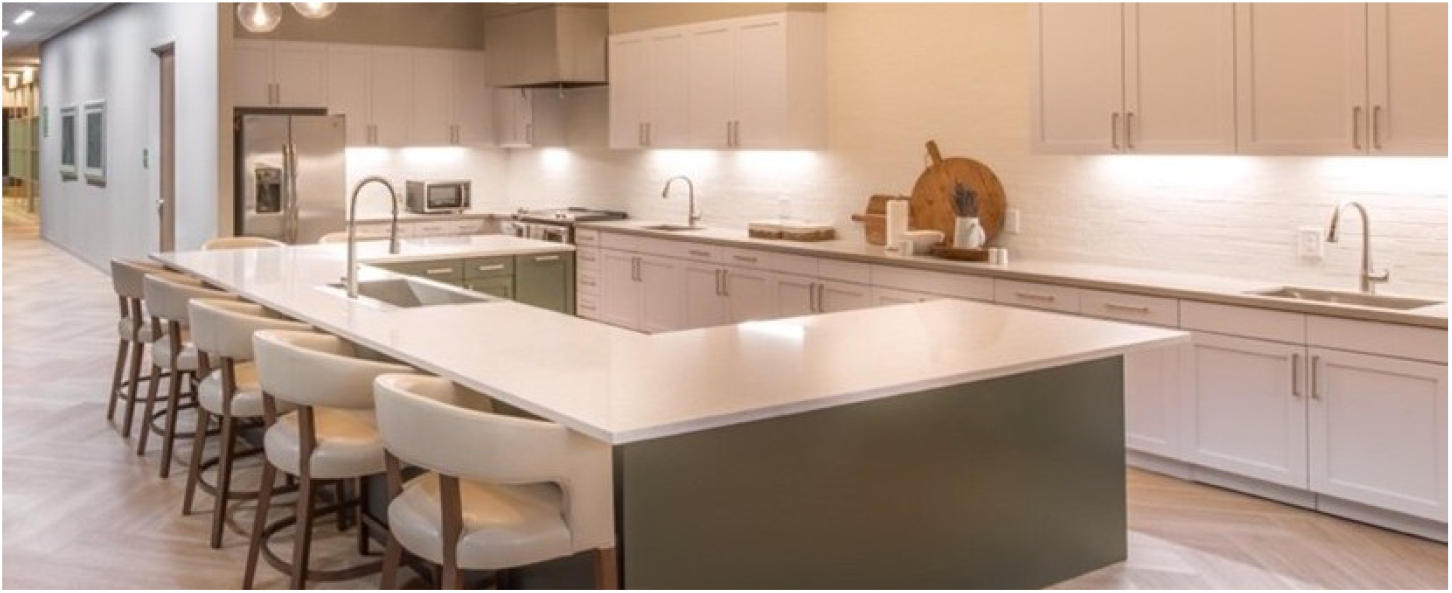
Kitchen Space

**Figure 2:**
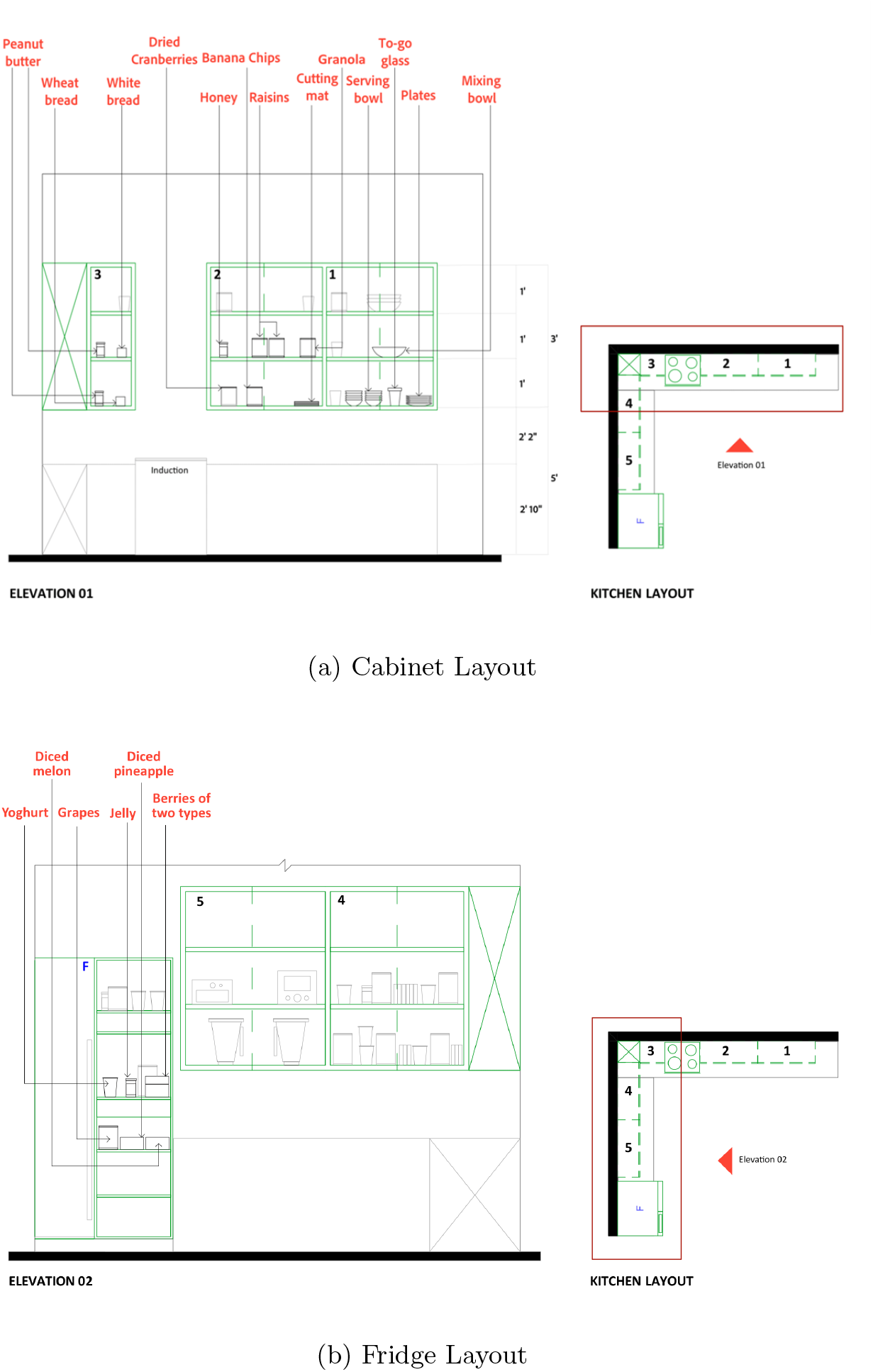
Standardized Spatial Configuration of Ingredients and Utensils

**Table 2:** 26 Steps of Yogurt Parfait Bowl Preparation Task.

| # | Description | Activity |
| --- | --- | --- |
| 1 | Take a bowl from the cabinet | Item Retrieval |
| 2 | Pick up the cooking mat from the cabinet. | Item Retrieval |
| 3 | Pick up the strawberry flavored yogurt from the refrigerator. | Item Retrieval |
| 4 | Scoop 4 spoons of yogurt into the bowl or glass. | Food Preparation |
| 5 | Return the yogurt to the refrigerator, and pick up the diced pineapples | Item Retrieval |
| 6 | Add a spoonful of diced pineapples on top of the yogurt in the bowl. | Food Preparation |
| 7 | Return the pineapples to the refrigerator and pick up the blue berries. | Item Retrieval |
| 8 | Add a spoonful of blue berries on top of the yogurt in the bowl. | Food Preparation |
| 9 | Return the blue berries to the refrigerator. | Item Retrieval |
| 10 | Pick up the banana chips in the cabinet. | Item Retrieval |
| 11 | Add a spoonful of banana chips on top of the yogurt in the bowl. | Food Preparation |
| 12 | Return the banana chips to the cabinet, pick up the raisins. | Item Retrieval |
| 13 | Add a spoonful of raisins on top of the yogurt in the bowl. | Food Preparation |
| 14 | Return the raisins to the cabinet and pick up the chocolate almonds. | Item Retrieval |
| 15 | Add a spoonful of chocolate almonds on top of the yogurt in the bowl. | Food Preparation |
| 16 | Return the chocolate almonds to the cabinet and pick up the granola. | Item Retrieval |
| 17 | Add a spoonful of granola on top of the yogurt in the bowl. | Food Preparation |
| 18 | Return the granola to the cabinet. | Item Retrieval |
| 19 | Grab a serving bowl and scoop 2 spoons of parfait into the serving bowl. | Food Preparation |
| 20 | Grab a to-go bowl and scoop remaining parfait into the to-go bowl. | Food Preparation |
| 21 | Pick up the honey in the cabinet. | Item Retrieval |
| 22 | Drizzle 1 tablespoon of honey on top. | Food Preparation |
| 23 | Return the honey to the cabinet. | Item Retrieval |
| 24 | Grab a to-go lid from the cabinet to cover the to-go bowl. | Item Retrieval |
| 25 | Place the serving bowl and to-go-bowl in the refrigerator. | Item Retrieval |
| 26 | Take the utensils to the sink (spoons, bowl, and mat). | Item Retrieval |

### 2.4 Data Acquisition and Preprocessing

#### 2.4.1 Wrist Sensor Data

Participants were equipped with tri-axial accelerometers (GENEActiv, ActivInsights Ltd) on both wrists, which were calibrated using manufacturer-provided software. Data were sampled at 100 Hz to capture high-resolution upper-limb motor activity during the 26-step yogurt preparation task. The wrist sensors recorded tri-axial acceleration (*Acc*_*x*_, *Acc*_*y*_, *Acc*_*z*_) and skin temperature via an integrated thermistor, as shown in Table 3. To quantify overall movement intensity independent of sensor orientation, we also derived the acceleration magnitude,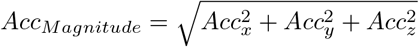. Wrist sensor data were successfully obtained from all 18 participants.

**Table 3:** Wrist Sensor Data.

| Data | Description |
| --- | --- |
| acceleration x, y, z | Raw tri-axial acceleration measurements (g) captured at 100 Hz. |
| thermistor | Skin temperature measurements ($^{\circ}\text{C}$) recorded from the wrist-worn thermistor to monitor physiological heat flux |
| acceleration magnitude | The vector magnitude calculated as $\sqrt{x^2 + y^2 + z^2}$, used to quantify overall physical activity intensity and movement displacement |

#### 2.4.2 Eye-Tracking Data

Visual attention and oculomotor behaviors were captured using Pupil Labs Core eye-tracking glasses (Pupil Labs GmbH) at a sampling rate of 30 Hz. The system generated two primary data streams: gaze position (17 features) and pupil position (16 features), as detailed in Table 4. Due to technical failures during collection, data from three participants in the MCI group were excluded, leaving a final eye-tracking dataset of 15 participants. From the raw coordinates, we derived 10 additional saccade features to characterize rapid eye movements [59]. For instance, the 3D saccade length for the eye center was calculated as, 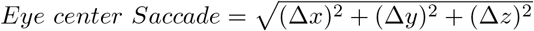, where Δ*x*, Δ*y*, and Δ*z* represent the coordinate changes between consecutive samples at the device’s sampling rate. These features, listed in Table 4, allow for the analysis of visuospatial navigation and potential delayed saccadic responses characteristic of cognitive impairment.

**Table 4:** Eye-Tracking Data.

| Feature | Description |
| --- | --- |
| **Gaze Features (17)** |  |
| gaze norm pos x, y (2) | Normalized x, y position of the gaze point in the world-camera coordinate system |
| gaze point 3d x, y, z (3) | x, y, z-coordinate of the 3D gaze point (point where the subject is looking) in the world-camera coordinate system |
| left eye center 3d x, y, z (3) | x, y, z-coordinate of the center of left eyeball in the world-camera coordinate system |
| right eye center 3d x, y, z (3) | x, y, z-coordinate of the center of right eyeball in the world-camera coordinate system |
| left gaze normal x, y, z (3) | x, y, z-component of the visual axis direction for left eye in the world-camera coordinate system |
| right gaze normal x, y, z (3) | x, y, z-component of the visual axis direction for right eye in the world-camera coordinate system |
| **Pupil Features (16)** |  |
| left pupil norm pos x, y (2) | Normalized x, y position of the left pupil in the eye image frame |
| right pupil norm pos x, y (2) | Normalized x, y position of the right pupil in the eye image frame |
| left ellipse center x, y (2) | x, y-coordinate of the left ellipse center (pupil) in image pixels |
| right ellipse center x, y (2) | x, y-coordinate of the right ellipse center (pupil) in image pixels |
| left ellipse axis a, b (2) | Length of the first, second axis of the left pupil ellipse in pixels |
| right ellipse axis a, b (2) | Length of the first, second axis of the right pupil ellipse in pixels |
| left ellipse angle | Angle of the left ellipse in degrees |
| right ellipse angle | Angle of the right ellipse in degrees |
| left pupil diameter | Diameter of the left pupil in image pixels |
| right pupil diameter | Diameter of the right pupil in image pixels |
| **Saccade Features (10)** |  |
| saccade gaze norm pos | Euclidean distance between consecutive normalized gaze positions |
| saccade gaze point 3d | Euclidean distance between consecutive 3D gaze point coordinates |
| saccade left eye center 3d | Euclidean distance between consecutive 3D coordinates of the left eye center |
| saccade right eye center 3d | Euclidean distance between consecutive 3D coordinates of the right eye center |
| saccade left pupil norm pos | Euclidean distance between consecutive normalized positions of the left pupil |
| saccade right pupil norm pos | Euclidean distance between consecutive normalized positions of the right pupil |
| saccade left pupil ellipse center | Euclidean distance between consecutive positions of the left pupil ellipse center |
| saccade right pupil ellipse center | Euclidean distance between consecutive positions of the right pupil ellipse center |
| saccade left gaze normal | Euclidean distance between consecutive vectors representing the visual axis direction for the left eye |
| saccade right gaze normal | Euclidean distance between consecutive vectors representing the visual axis direction for the right eye |

### 2.5 Multimodal Data Analysis Pipeline

Our analysis pipeline, shown in Figure 3, follows the standard human activity recognition pipeline, including sliding window-based segmentation, feature extraction, and classification with machine learning models [60]. For window-level classification, we used two different approaches: (1) shallow machine learning models and (2) deep learning models. The multimodal time-series from wrist sensors and eye-tracking data were first segmented using a 10-second sliding window, namely analysis window, with 50% overlap. This window size was selected to capture sufficient length of activity context in meal preparation, including sporadic and longer-duration behaviors shown in Table 2 [61, 62]. For shallow machine learning models, we utilized the TSFRESH package [63] to extract time-series statistical features from each window, including mean, variance, mean absolute change (MAC), root mean square (RMS), zero crossing rate (ZCR), and the empirical cumulative distribution function (ECDF) with five components [64, 65, 63]. These features were selected to preserve arbitrary sensor distribution characteristics and have demonstrated high classification performance in wearable sensor-based activity recognition [66, 67]. For multimodal data analysis, when using shallow machine learning models, like Random Forest (RF), we first concatenated modality-specific features from the left wrist, right wrist, and eye-tracking sensors into a single feature vector, which is then processed by the classifier. For deep learning-based analysis, like the Deep Convolutional LSTM (DeepConvLSTM) model [68], the raw time-series data from the analysis window, were directly processed with deep learning models to learn multimodal feature representations. To align the sampling rates, wrist sensor data were downsampled to 30 Hz to match the eye-tracking rate, and all channels were concatenated along the channel axis.

**Figure 3:**
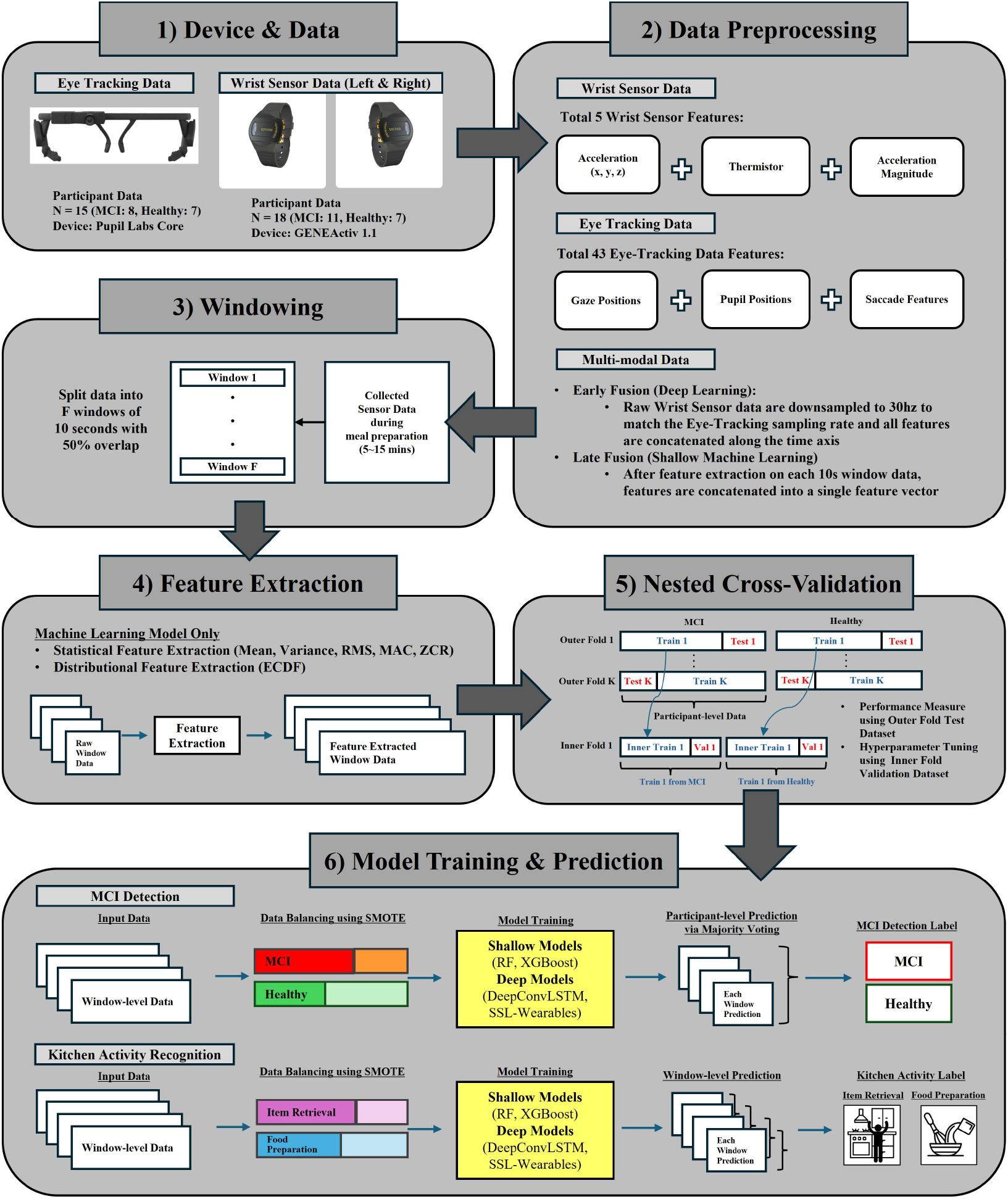
Overview of the analysis and evaluation framework for MCI Detection and Kitchen Activity Recognition using multimodal wearable sensor data

### 2.6 Machine Learning Models

#### 2.6.1 Shallow Machine Learning

We used RF and eXtreme Gradient Boosting (XGBoost), which were successfully used in various HAR tasks [69, 70].

#### 2.6.2 Deep Learning Model

Following the best practice in HAR, we explore two ways of using deep learning models for our analysis: (1) training from scratch and (2) fine-tuning models pretrained with large-scale datasets. For training a model from scratch, we used the DeepConvLSTM model [68], which was successful in various HAR tasks as a baseline deep learning model for benchmarking [70]. We also selected DeepConvLSTM for its compact representation, considering the small dataset in this feasibility study. The architecture consists of four stacked 2D convolutional layers with batch normalization and ReLU activation, followed by a bidirectional LSTM and a fully connected output layer. For fine-tuning the pretrained model, we primarily used the model for wrist sensor data. Specifically, we used *SSL-Wearables* [71], a ResNet-based architecture pretrained on the UK Biobank accelerometer dataset [72], comprising over one week of accelerometer recordings from 100,000 participants using a self-supervised learning framework. This model was successfully adapted to various benchmark tasks in HAR, including the clinical population (older adults with Parkinson’s Disease). To match the pretrained model’s expected input format, we resampled our 100 Hz wrist accelerometer data to 30 Hz using linear interpolation, resulting in 300 samples per 10-second window. Only the three-axis acceleration channels (Acc_*x*_, Acc_*y*_, Acc_*z*_) were used as input to maintain compatibility with the pretrained weights. For fine-tuning the *SSL-Wearables* model for our tasks, we froze the pretrained ResNet backbone and attached a multilayer perceptron (MLP) classifier head. The MLP consists of two hidden layers with 512 and 256 units, respectively, each followed by ReLU activation and dropout (rate = 0.3), and a final output layer. For the eye-tracking signal, we didn’t use the pretrained model. While pretrained frameworks, such as the Oculomotor Behavior Framework (OBF) [73], exist for extracting feature representations from gaze scanpaths, they were primarily optimized for stationary, desktop-mounted eye-trackers and validated on tasks like stimulus prediction or clinical screening (e.g., autism spectrum disorder). We considered the OBF’s pretrained task irrelevant to our study, which uses portable eye-tracking glasses during dynamic activities in a kitchen environment. Furthermore, a pretrained model specifically designed to capture eye-movement dynamics for HAR is currently absent in the literature.

### 2.7 Experiments and Evaluation Protocol

#### 2.7.1 Recognition Tasks

Our multimodal wearable analysis pipeline was evaluated for two different tasks: 1) MCI detection and 2) Kitchen Activity Recognition (KAR). For MCI detection, all windows from each participant are labeled as NC or MCI according to the participant’s diagnosis. The window-level prediction was aggregated via majority voting for participant-level classification. In addition to participant-level MCI detection analysis, we also reported the F1 scores for each activity class (*item retrieval* or *food preparation*), to understand which activity was most associated with cognitive decline. For the KAR task, we labeled each window with the activity label of either *item retrieval* or *food preparation* at the final timestep of the window, enabling the model to immediately detect the onset and offset of activities via sliding-window analysis. For both MCI detection and the KAR task, the performance was evaluated using 10 runs of participant-wise nested 5-fold cross-validation, with training, validation, and test participant splits of 60%, 20%, and 20%, respectively. We used the F1 score to assess model performance and a 95% Wilson score confidence interval [74] to assess statistical significance. To address class imbalance, we applied the Synthetic Minority Over-sampling Technique (SMOTE) [75] with *k* = 5 neighbors for all models. Hyperparameter optimization was performed using Optuna [76] with

Tree-structured Parzen Estimator (TPE) sampling. For RF, we optimized `max_depth` ∈ {3, 5, 7, 10}, `min_samples split` ∈ {2, 5, 10}, and n_estimators ∈ {100, 200, 500}. For XGBoost, we optimized `n_estimators` ∈ {100, 200, 500}, `learning_rate` ∈ {0.01, 0.1, 0.3}, and `max_depth` ∈{3, 5, 7, 10 }. For DeepConvLSTM, we optimized the following hyperparameters: number of LSTM units ∈{32, 64, 128}, number of LSTM layers ∈{1, 2, 3}, convolutional filter size ∈{3, 5, 7}, number of filters {32, 64, 128}, number of convolutional layers ∈{2, 3, 4}, dropout rate ∈ [0.1, 0.5], and learning rate 10^*−*4^, 10^*−*5^ . Training was performed using the Adam optimizer with early stopping (patience of 10 epochs) and a maximum of 50 epochs, selecting the model with the lowest validation loss. For the pretrained model, we froze the pretrained ResNet backbone, and the MLP classifier was trained using Adam optimizer with learning rate 10^*−*3^ and weight decay 10^*−*4^, employing early stopping with patience of 10 epochs and a maximum of 50 epochs.

#### 2.7.2 Feature Importance Analysis

We identified the most discriminative features for MCI detection and KAR tasks. For shallow machine learning models (RF and XGBoost), we used the impurity-based (Mean Decrease in Impurity) feature importance, which measures the total reduction in Gini impurity contributed by each feature across all trees in the ensemble [77]. For the deep learning models, we adopted SHAP (SHapley Additive exPlanations) [78] to compute feature attributions. Specifically, we employed the DeepExplainer algorithm, which leverages DeepLIFT-based approximations of Shapley values optimized for deep neural networks, rather than model-agnostic ensemble methods such as KernelSHAP, enabling efficient and accurate attribution for our convolutional-recurrent architecture. The SHAP values were aggregated across the temporal dimension to obtain per-feature importance scores, and mean absolute SHAP values were computed across all test samples and cross-validation folds to ensure statistically significant importance estimates.

## 3 Results

The experimental results are based on data from 18 participants (11 MCI, 7 NC), with specific sample sizes varying by modality. Due to technical issues encountered during the collection of eye-tracking data (Sec. 2.4.2), the analysis for eye-tracking and multi-modal models was limited to 15 participants (8 MCI, 7 NC), while the wrist-sensor analysis utilized the complete dataset of 18.

### 3.1 MCI Detection

The performance of MCI detection across different modalities and model architectures is summarized in Table 5a. When using wrist sensor data only, the *SSL-Wearable*s model achieved the highest F1 score of 0.759. When using eye-tracking data and multimodal data, the DeepConvLSTM model achieved the highest F1 score of 0.749 and 0.757, respectively. When detecting MCI within each activity context (Table 5b), DeepConvLSTM using eye-tracking data showed the best performance with 0.762 F1 score during the *item retrieval* activity. For detecting MCI during *food preparation*, RF with multimodal sensors showed the highest F1 score with 0.701 F1 score. Figure 4a shows the feature importance analysis for MCI detection when using multimodal data analysis with the DeepConvLSTM model, which performed the best in Table 5a.

**Table 5:** Performance for MCI Detection. **Bolded** numbers are the best performing model for each row.

(a) Performance across all kitchen activities. **Highlighted** number is the best performing model.
| Modality | RF | XGBoost | DeepConvLSTM | SSL-Wearables |
| --- | --- | --- | --- | --- |
| Wrist Sensor Data | 0.637 ± 0.067 | 0.678 ± 0.063 | 0.573 ± 0.067 | **0.759 ± 0.045** |
| Eye-Tracking Data | 0.723 ± 0.100 | 0.693 ± 0.110 | **0.749 ± 0.101** | - |
| Multi-modal Data | 0.674 ± 0.102 | 0.692 ± 0.108 | **0.757 ± 0.101** | - |

| Activity Class | RF | XGBoost | DeepConvLSTM |
| --- | --- | --- | --- |
| **Wrist Sensor Data** |  |  |  |
| Item Retrieval | **0.669 ± 0.018** | 0.660 ± 0.019 | 0.598 ± 0.019 |
| Food Preparation | **0.584 ± 0.026** | 0.560 ± 0.026 | 0.516 ± 0.026 |
| **Eye-Tracking Data** |  |  |  |
| Item Retrieval | 0.753 ± 0.028 | 0.707 ± 0.031 | **0.762 ± 0.028** |
| Food Preparation | 0.655 ± 0.041 | 0.629 ± 0.046 | **0.671 ± 0.041** |
| **Multi-modal Data** |  |  |  |
| Item Retrieval | 0.683 ± 0.025 | 0.621 ± 0.027 | **0.732 ± 0.030** |
| Food Preparation | **0.701 ± 0.031** | 0.635 ± 0.034 | 0.641 ± 0.044 |

**Figure 4:**
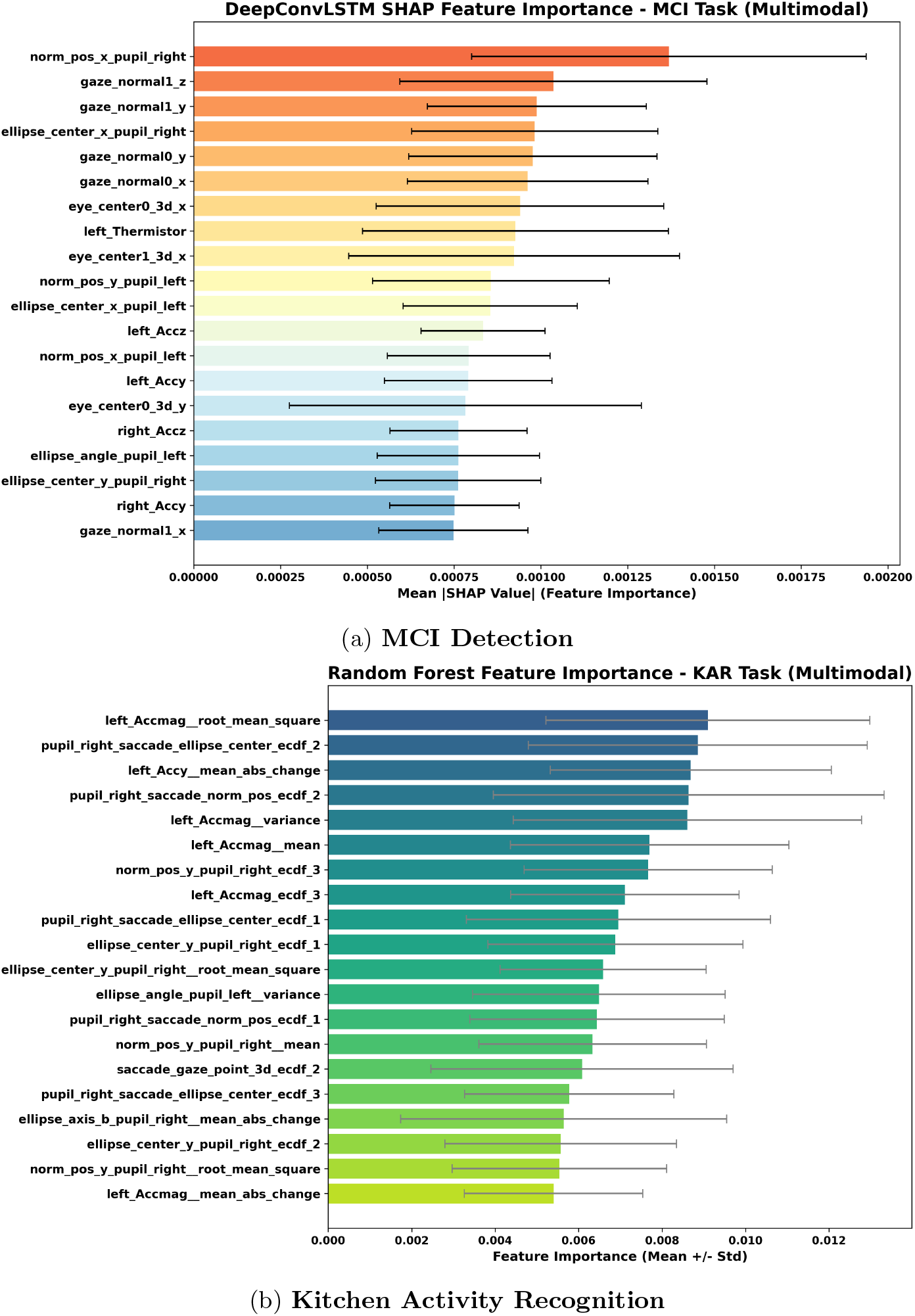
Feature importance analysis for multimodal features

### 3.2 Kitchen Activity Recognition

Table 6 present the F1scores for the classification of *item retrieval* and *food preparation* activities. For recognizing the *item retrieval* activity, the RF model with multimodal data achieved the best performance, with an F1 score of 0.672, and for recognizing the *food preparation* class, the *SSL-Wearables* model achieved the best performance, with an F1 score of 0.55. For both activities combined, the RF model with multimodal data achieved the best performance, with an F1 score of 0.596. Figure 4b shows the feature importance analysis for the KAR task when using multimodal data with the RF model, which was the best-performing model overall from Table 6.

**Table 6:**
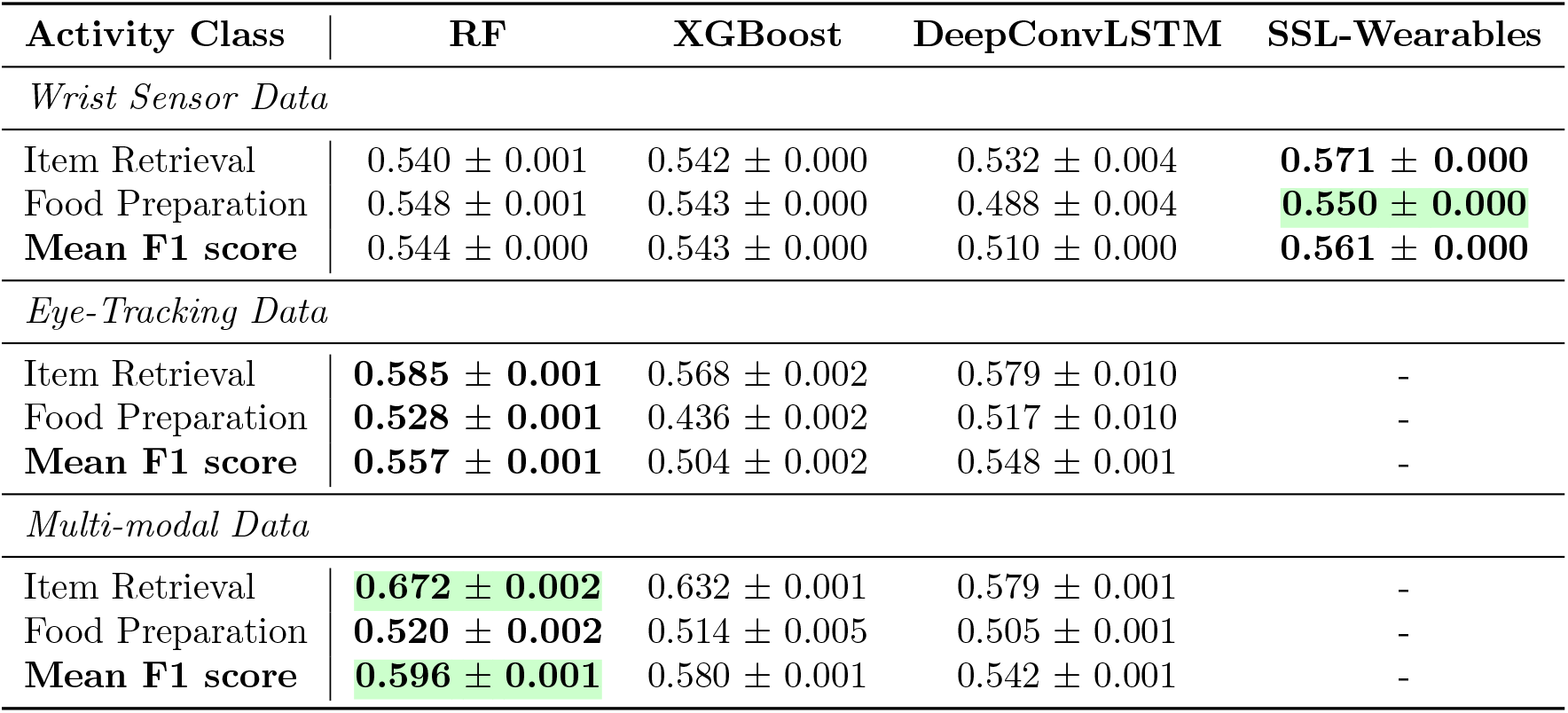
Performances for Kitchen Activity Recognition. Bolded numbers are the best performing model for each row. Highlighted numbers are the best performing models for recognizing Item Retrieval-only, Food Preparation-only, and combining both on average.

| Activity Class | RF | XGBoost | DeepConvLSTM | SSL-Wearables |
| --- | --- | --- | --- | --- |
| *Wrist Sensor Data* |  |  |  |  |
| Item Retrieval | 0.540 ± 0.001 | 0.542 ± 0.000 | 0.532 ± 0.004 | **0.571 ± 0.000** |
| Food Preparation | 0.548 ± 0.001 | 0.543 ± 0.000 | 0.488 ± 0.004 | **0.550 ± 0.000** |
| **Mean F1 score** | 0.544 ± 0.000 | 0.543 ± 0.000 | 0.510 ± 0.000 | **0.561 ± 0.000** |
| *Eye-Tracking Data* |  |  |  |  |
| Item Retrieval | **0.585 ± 0.001** | 0.568 ± 0.002 | 0.579 ± 0.010 | - |
| Food Preparation | **0.528 ± 0.001** | 0.436 ± 0.002 | 0.517 ± 0.010 | - |
| **Mean F1 score** | **0.557 ± 0.001** | 0.504 ± 0.002 | 0.548 ± 0.001 | - |
| *Multi-modal Data* |  |  |  |  |
| Item Retrieval | **0.672 ± 0.002** | 0.632 ± 0.001 | 0.579 ± 0.001 | - |
| Food Preparation | **0.520 ± 0.002** | 0.514 ± 0.005 | 0.505 ± 0.001 | - |
| **Mean F1 score** | **0.596 ± 0.001** | 0.580 ± 0.001 | 0.542 ± 0.001 | - |

## 4 Discussion

Our study introduces the first multimodal dataset of kitchen activities captured from 18 age-matched older adults (N=11 MCI, N=7 NC) in a semi-naturalistic kitchen environment. Our results demonstrated that the HAR-based screening system could identify MCI through complex behavioral patterns during kitchen activities, as well as physical and physiological features obtained from wearable devices, such as movement intensity and saccadic dynamics, as objective markers for cognitive decline.

### 4.1 MCI Detection

The *SSL-Wearable* model, utilizing only wrist sensor data, achieved an F1 score of 0.759, which is comparable to the performance of the DeepConvLSTM model using eye-tracking (0.749) and multi-modal datasets (0.757). This result is particularly noteworthy given that the DeepConvLSTM model reached only a 0.573 F1 score when restricted to wrist sensor data. Such a disparity demonstrates that the *SSL-Wearable* model, pre-trained on large-scale human activity datasets, learns generalizable feature representations that can be effectively fine-tuned for specialized tasks, such as monitoring kitchen activities in individuals with MCI. Compared to the wrist sensor data, eye-tracking data provided the most discriminative features for MCI detection Figure 4a. Specifically, gaze normal features (visual axis direction) and eye center features (3D spatial coordinates) emerged as the most critical variables. The robust performance of models incorporating these oculomotor features underscores their significant contribution to quantifying cognitive decline, suggesting that while wrist sensors capture functional motor patterns, eye-tracking provides essential insights into the underlying cognitive load and visuospatial difficulties. Furthermore, this success hints that developing foundation models for eye-tracking data, similar to those for wrist sensor data, could further enhance detection capabilities in daily living settings.

Figure 5 shows a direct comparison of accelerometer and eyetracking features for MCI and NC groups on average. Older adults with NC exhibited a notably higher mean acceleration along the x-axis and demonstrated significantly greater variance and Mean Absolute Change (MAC) across all tri-axial components compared to the MCI group. While thermistor-related features were identified as the 8th most influential feature in Figure 4a, the exploratory data analysis revealed that group-level differences in skin temperature were negligible compared to the more pronounced signatures in physical motor dynamics (Figure 5b). These findings suggest that cognitive decline is associated with reductions in upper-limb motor intensity and complexity during naturalistic kitchen activities. Our observations align with previous research investigating the link between psychomotor function and cognitive status [79], which reported diminished motor performance in tasks such as trail making and spiral tracing among individuals with MCI or ADRD [80, 81, 82].

**Figure 5:**
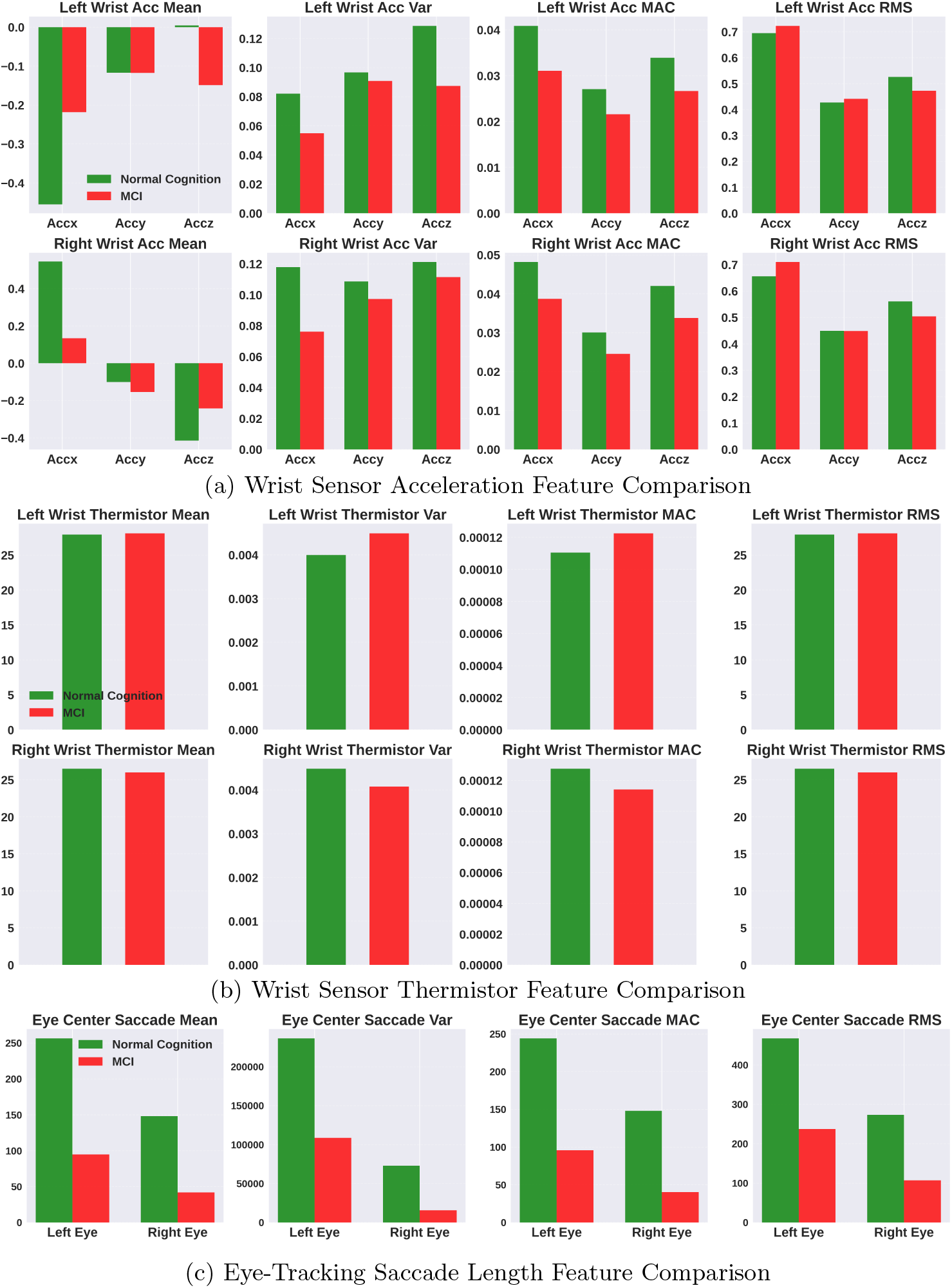
Comparison of wrist sensor and eye-tracking features between MCI and NC

Additionally, all saccade-length metrics in the MCI group were significantly lower than those in the NC group, indicating reduced saccadic eye-movement amplitude and dynamics. This observation aligns with established literature reporting prolonged latencies and diminished saccadic velocity in individuals with MCI, where such oculomotor abnormalities are closely correlated with the severity of cognitive impairment [83, 84, 85, 86, 87]. Beyond reactive saccades, the MCI group generates fewer anticipatory saccades, which are eye movements initiated before sufficient visual processing occurs, driven not by direct visual input but by cognitive mechanisms such as the prediction of next actions to be taken during the meal preparation tasks [88]. Previous studies reported that reductions in anticipatory saccades toward expected waypoints, along with impaired performance on antisaccade tasks, suggest deficits in inhibitory control and disrupted neural mechanisms involved in predictive processing [83, 89, 90].

Table 5b shows that the *Item Retrieval* period serves as a significantly more predictive of MCI than the *Food Preparation* phase. Specifically, the eye-tracking modality achieved its peak performance during item retrieval with an F1-score of 0.762 using the DeepConvLSTM model, which performance dropped to 0.671 during the preparation phase. This aligns with Bilau *et al*. [9], who reported that navigating cabinets and refrigerators poses a substantial cognitive challenge for individuals with MCI due to impaired spatial navigation and elevated cognitive load. This heightened cognitive demand is likely exacerbated by the specific working memory requirements of the retrieval task. Unlike the *Food Preparation* phase, where participants can frequently reference instructions kept directly in front of them, the *Item Retrieval* requires them to move away from the counter and leave the written protocol behind. They must internally maintain the identity and intended location of the item while simultaneously performing a visual search. Furthermore, the high model performance captured during retrieval tasks may be attributed to the complex oculomotor dynamics required for vertical scanning of multi-layer kitchen storage. Zhao *et al*. [83] demonstrated that vertical saccadic tasks exhibit higher sensitivity in detecting cognitive impairment compared to horizontal tasks, as vertical eye movements are more demanding of neurocognitive control mechanisms. Consequently, the *Item Retrieval* phase, which inherently integrates high-level cognitive planning with vertical oculomotor dynamics, has the potential for a robust digital marker for identifying early-stage cognitive decline in naturalistic environments.

### 4.2 Kitchen Activity Recognition

The superior performance of the multi-modal configuration, shown in Table 6, underscores the complementary nature of physical motion and oculomotor signals in characterizing complex kitchen activities. Feature importance analysis in Figure 4b identified the left wrist acceleration (X-axis) and acceleration magnitude-based statistical features as the most discriminative variables along with other eye-tracking features. This finding is consistent with established HAR literature, which designates tri-axial acceleration as a primary digital biomarker for distinguishing fine-grained motor patterns [71]. Notably, the RF model exhibited better classification performance than the DeepConvLSTM architecture across most tasks, likely due to the inherent challenges of training deep learning models on a small dataset for complex activity recognition tasks, which also makes it difficult to identify strict temporal boundaries between activities. Similarly, from MCI detection, fine-tuning *SSL-Wearables* model outperformed the DeepConvLSTM model and showed the best performance for recognizing the *Food Preparation* task, demonstrating the effectiveness of adapting a pretrained model from a large-scale dataset.

Figure 6 shows the comparison of wrist sensor and eye-tracking signals for each kitchen activity class for the NC and MCI groups. Acceleration intensities across all axes were notably higher during the *Item Retrieval* period compared to the *Food Preparation* phase (Figure 6a). This suggests a greater intensity of upper limb motor function during retrieval, which requires reaching, grasping, and transporting objects, as opposed to the more localized movements involved in food preparation. Interestingly, while the NC group showed a clear difference in acceleration intensity between the two activities, the MCI group exhibited only marginal differences. Eye center saccade analysis also showed that saccade variance and Mean Absolute Change (MAC) were generally higher during food preparation for the NC group, whereas values remained relatively consistent across activities for the MCI cohort (Figure 6b). Consistent with our MCI detection findings, the NC group demonstrated significantly higher saccadic dynamics than the MCI group across both activity contexts. These results suggest that while healthy older adults adapt their motor and visual strategies to the specific demands of the task, individuals with MCI may lack this behavioral flexibility, resulting in a more uniform and diminished physiological signature across complex daily living activities.

**Figure 6:**
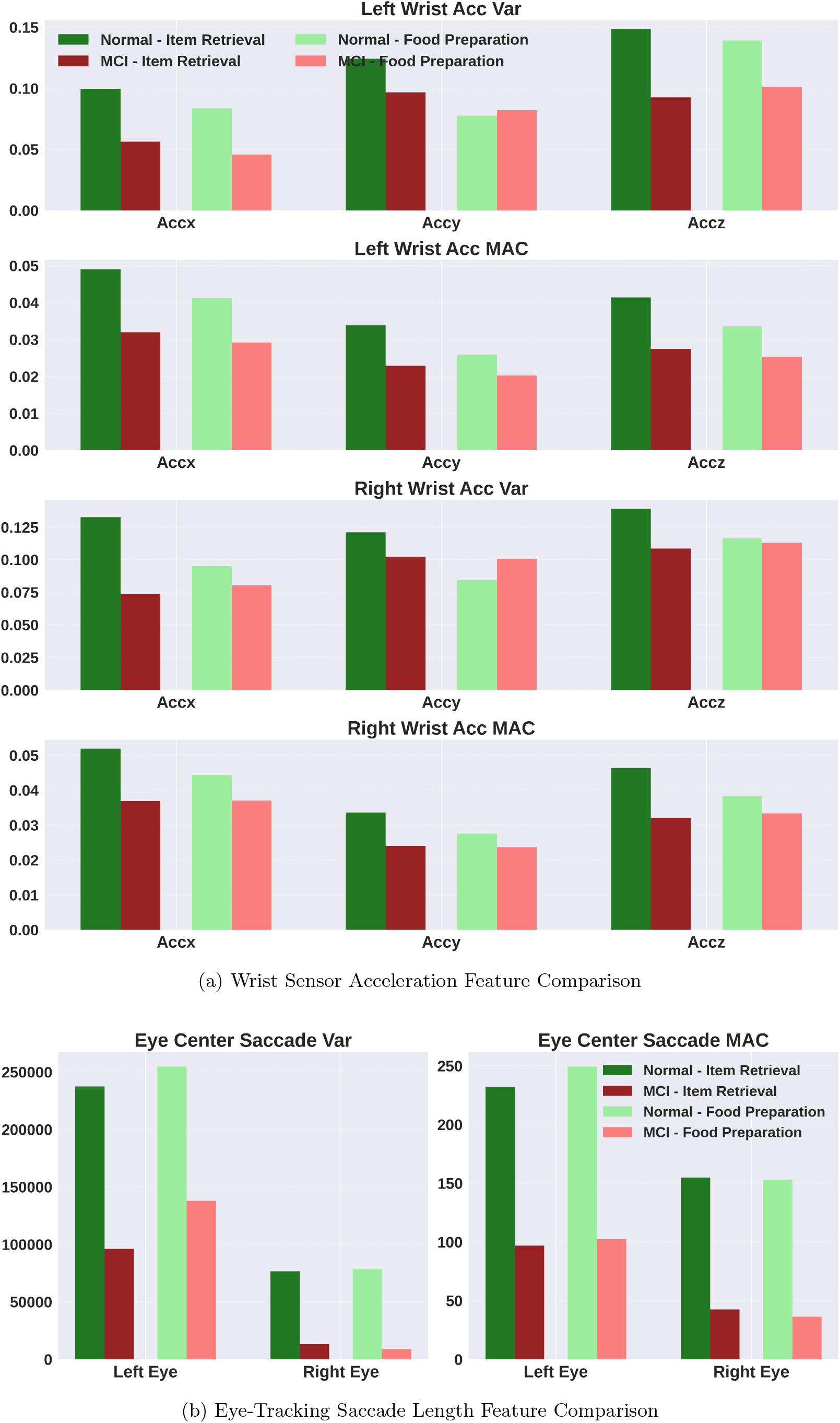
Comparison of wrist sensor and eye-tracking features between MCI and NC in *Item Retrieval* and *Food Preparation* activities.

### 4.3 Limitations and Future Work

This study demonstrates the feasibility of quantifying cognitive decline using multimodal wearable sensors in the kitchen environment. In our experimental design, all participants performed meal preparation tasks under standardized conditions with identical recipes and fixed locations for ingredients and utensil. However, kitchen environments and designs vary considerably in real-world settings, as do approaches to food preparation. To effectively support aging-in-place and functional independence in kitchen activities, an investigation of actual behaviors in participants’ home environments is essential. Additionally, MCI presents significant heterogeneity that necessitates large-scale subject recruitment in future work to validate the generalizability of our findings and address potential biases associated with demographic variables, such as age, sex, race, and education levels. Also, the inclusion of participants across a diverse spectrum of cognitive impairment, from MCI to mild ADRD, is essential to characterize progressive changes in motor function and visual attention during kitchen activities. Moreover, our study in Table 5b shows the importance of behavior markers based on activity contexts, and this calls for further research integrating advanced techniques on human activity recognition and MCI detection as a holistic framework for behavior analysis in the kitchen. Our ongoing work includes ambient video recordings or ego-centric camera views for kitchen activity analysis to further capture environmental cues associated with activity context and cognitive impairments [52, 91]. The potential of such vision-based integration is exemplified by CHEF-VL [53], which demonstrates how ambient video can effectively monitor environmental state transitions, such as appliance status or object placement, to refine action recognition and detect sequencing deviations. For example, our participants with MCI reported that it was more difficult to search for objects above their eye level or located in the far reaches of the cabinet, which can be captured with additional ambient and wearable cameras. Furthermore, we envision integrating activity-aware ambient guidance systems with smart earbuds to capture acoustic environmental cues and head-motion dynamics, thereby enriching the behavioral modeling required to provide real-time assistance for individuals with cognitive decline [92].

## 5 Conclusion

Detecting MCI remains a significant challenge due to its subtle behavioral manifestations and requires non-invasive, scalable screening methods. Previous studies have primarily focused on quantifying mobility impairments, like gait, in controlled environments, and very few have examined behavioral markers in other activities of daily living related to cognitive decline. Our study addresses this gap by using wrist and eye-tracking wearable sensors to capture fine-grained motor and visuospatial behaviors during kitchen activities, achieving a 76% F1 score in distinguishing MCI from NC. This work not only highlights the feasibility of assessing cognitive functioning through routine activities but also underscores the potential for continuous, unobtrusive monitoring in real-world settings. Our findings suggest that integrating daily activities into behavioral sensing could redefine early intervention strategies and offer potential for therapeutic guidance in MCI, moving towards a more proactive and accessible model of cognitive health monitoring.

## Data Availability

All data produced in the present study are available upon reasonable request to the authors

## Acknowledgments

Hyeokhyen Kwon is partially supported by the National Institute on Deafness and Other Communication Disorders (Grant 1R21DC021029-01A1) and NCNM4R 2024-2025 Pilot Project Grants (AWD-006196-G1). The Charlie and Harriet Shaffer Cognitive Empowerment Program is supported by a generous investment from the James M. Cox Foundation and Cox Enterprises, Inc. We thank Jennifer DuBose, Zhi Tan, Bolaji Omofojoye, Alexander Adams, Katie Schreiber, Elahn Little, Madhuparna Sastakar, Abdurrahman Baru, and Marwan Shagar for their invaluable contributions to this study. We also thank the members, care partners, and staff at the Charlie and Harriet Shaffer Cognitive Empowerment Program for participating in this study and supporting our research. We also thank our colleagues and staff at the Georgia Institute of Technology, Emory University, and Northeastern University for their invaluable partnership.

